# Closed form solution of the SIR model for the COVID-19 outbreak in Italy

**DOI:** 10.1101/2020.06.06.20124313

**Authors:** Riccardo Giubilei

## Abstract

The CODIV-19 outbreak in early 2020 generated a tremendous effort of epidemiologists and researchers to fit the experimental data with the solutions of the SIR model equations [1] or with more sophisticated models. In this paper we show that under same hypotheses, a closed form solution exists that reasonably fits the experimental data for Italy, and the results can be extended to any other area.

## Introduction

The COVID-19 epidemic in 2020 has aroused the interest of epidemiologists and researchers with the aim to validate the most common models that describe the evolution of a epidemic and to provide a reliable forecast of the epidemic dynamics ([2][3][4][5]).

Since the beginning of the outbreak, first in China and Far East, then in Italy and Europe, a lot of information was available on the Web [6]. Normally data about total infected, actual infected, recovered, dead are reported daily ([7][8] for Italy) and allow to carry out analyses to verify the fit of same model with experimental data.

The following figure 1 refers to Italy [7] and reports the daily new infected, from the 20^th^ of February to mid-May. Data are affected by statistical fluctuations and by systematic biases that make them difficult to be interpreted. It is noted, for instance, that there is a sinusoidal behavior of the curve, around a mean value, with a period 7 days, that can be due to test results accumulated for a few days and accounted the days after.

**Figure 1:**
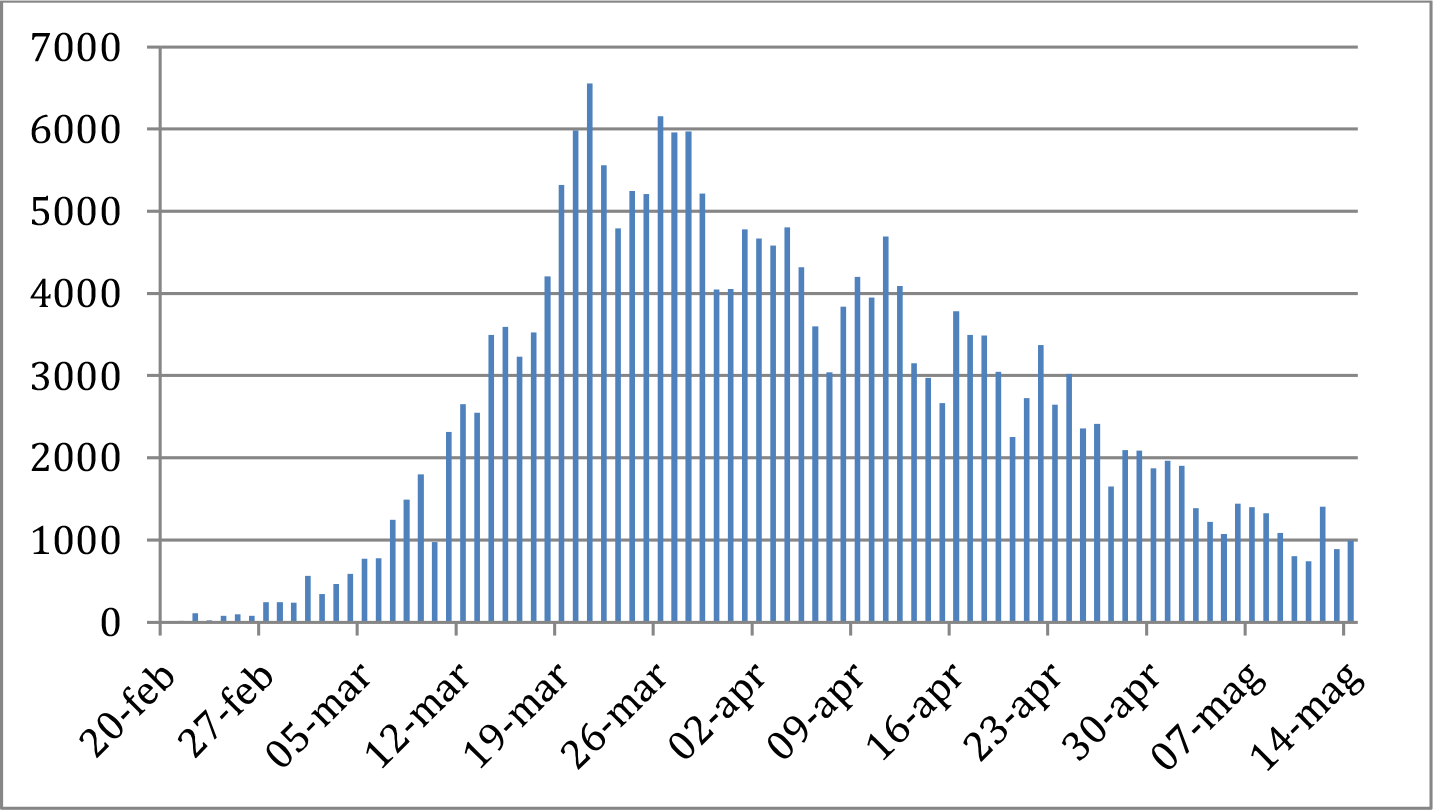
Daily new infected in Italy [7].

An interesting point is to find a curve that fits these experimental data. Among several functions considered, a very good fit has been obtained by using a Gumbel probability density function (pdf)[9], given by:

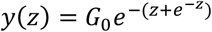

Where 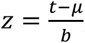, being μ the mode of the function, that identifies its maximum. The question was to understand whether this “double exponential” function is simply a good fit of the experimental data or it is a good fit because this type of functions are the solution of the differential equations that govern the dynamics of the epidemic evolution.

The model taken for the investigation is a compartmental model called SIR model, proposed by Kermack and McKendrick in 1927 ([10],[11]). The model consists of three compartments: “S” for the number of susceptible, “I” for the number of infectious, and “R” for the number of removed (recovered or deceased) individuals.

The dynamics of the epidemic according to this model is a set of non-linear differential equations involving S(t), I(t) and R(t) and a closed form solution is not trivial ([12], [13]). In the following we make some assumptions (later on verified through the experimental data) and on the basis of these assumptions we find a very simple closed form solution, that fits the experimental data.

## The SIR model

The SIR model can be expressed by the following set of ordinary differential equations [1]:

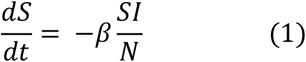

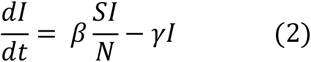

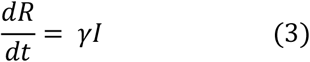

where *S*(*t*) is the susceptible population, *I*(*t*) is the infected population and *R*(*t*) is the removed population (either by death or recovery), and *N = S*(*t*)*+ I*(*t*) *+ R*(*t*). N is constant, being the model without vital dynamics (birth and death).

Normally *S*(*t*) decreases and *R*(*t*) increases significantly, until the herd immunity is achieved. In the case of COVID-19 the social distancing has been put in place immediately and the epidemic slowed down in few weeks.

*S*(*t*), starting from an initial value S_0_, is reduced by less than 0,06% worldwide and 0,4% in Italy [7], one of the most affected countries. On this ground we make the assumption that *S/N* ≅ 1, therefore the second SIR equation becomes:

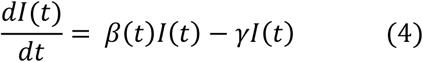

in which we assume that *γ*, the removal rate, is nearly constant and *β*(t), the transmission rate, is a function of time, varying according to the social distancing measures put in place. Equation (4) is a linear differential equation of the first order, whose solution, for *I*(0) = *I*_0_ is[14]:

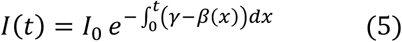

Let us assume that *β*(*t*) has an exponential shape, i.e. *β*(*t*) = *β*_0_*e*^*−αt*^. This conjecture will be verified by fitting experimental data. The exponent of the previous equation becomes:

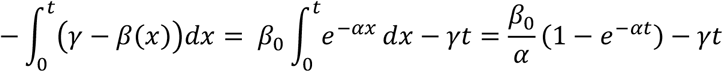

and finally:

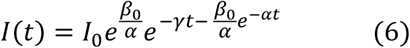

The function is of the “double exponential” type and it looks like the Gumbel pdf, i.e. 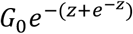.We define now:

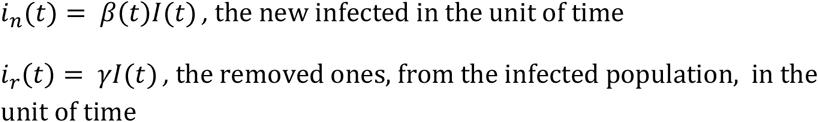

then:

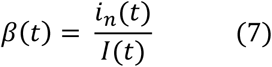

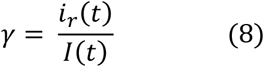

The previous equations can be used to estimate *β*(*t*) and *γ* from the experimental data. We take the data relevant to Italy [7]. The t=0 corresponds to the 20^th^ of February 2020. The unit of time is the day.

In the following diagram we show *β*(*t*) calculated accordingly to (7), and based on the experimental data for *i*_*n*_(*t*), the new daily infected, and I(*t*), the active cases, found in [7]. Experimental data in fig. 2 confirms that *β*(*t*) has a negative exponential behavior, thus validating the conjecture leading to Equation (6).

**Figure 2:**
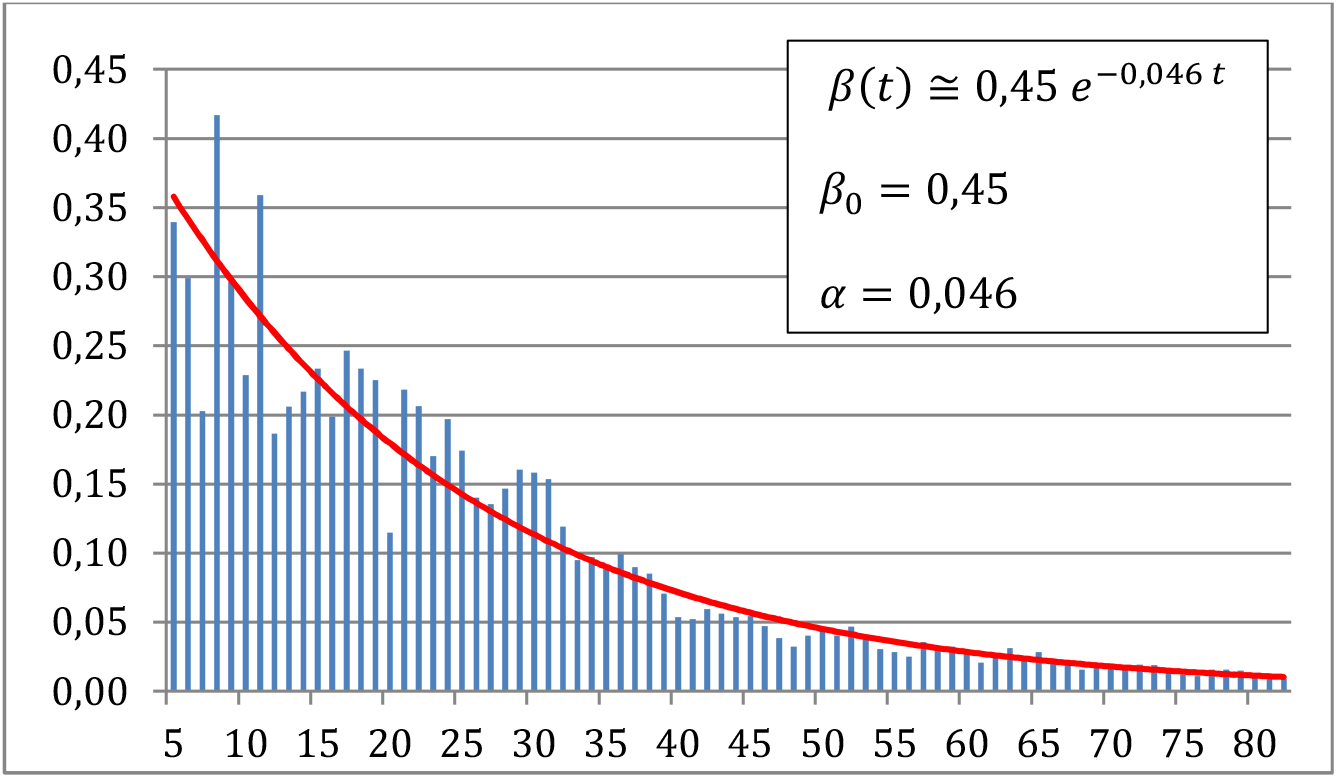
estimation of *β*(*t*) – x-axis: number of days from the 20^th^ of February.

The *γ* parameter is not constant. Also averaging the daily value over 7 days, its moving average lies between 0,04 and 0,02 day^-1^. We decided to retain a constant value, then *γ* ≅ 0,03 day^-1^.

Now *β*_0_, a and γ are estimated and we can use Equation (6) to fit the experimental data for I(t), trying to tune the *I*_0_ initial value. Figure 4 shows the results for *I*_0_ = 65.

**Figure 3:**
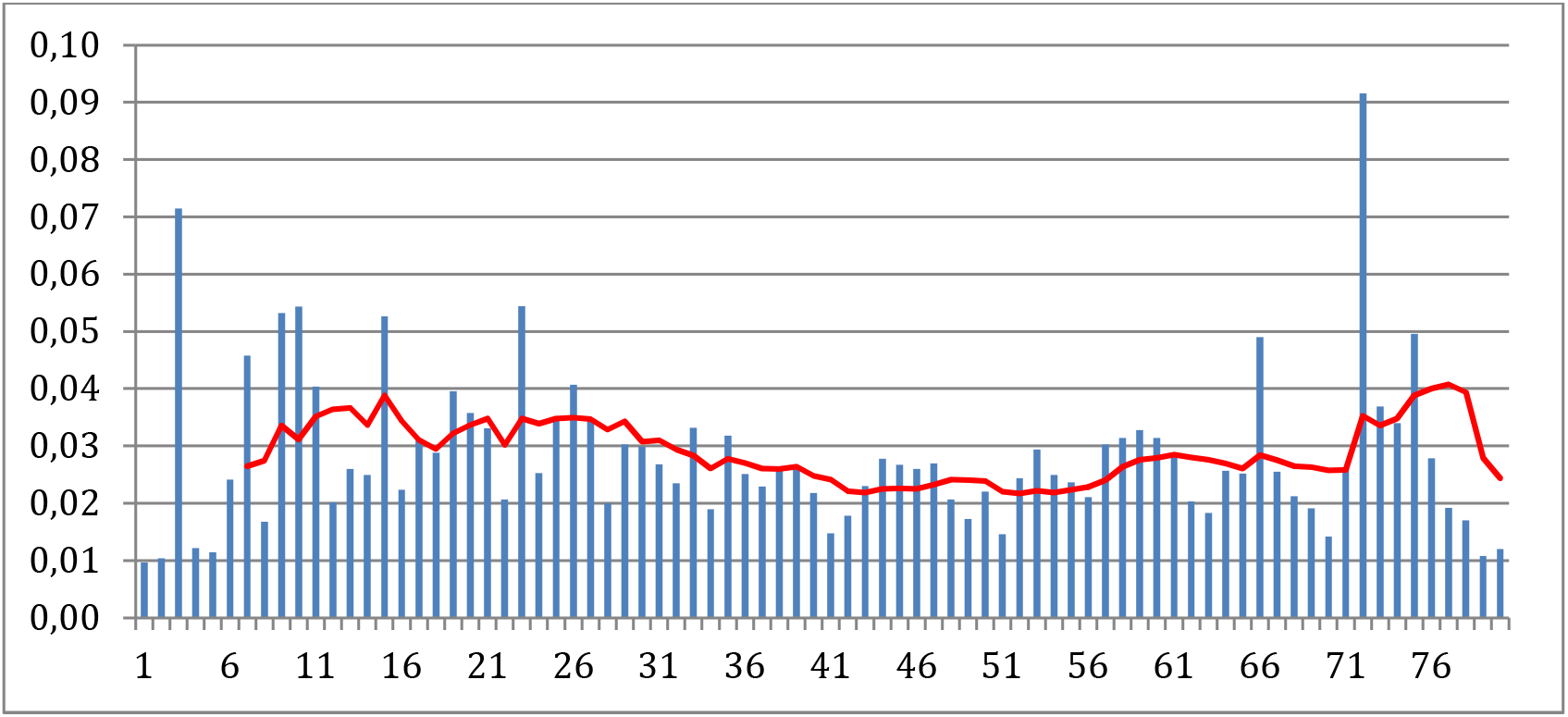
estimation of *γ* – x-axis: number of days from the 20^th^ of February.

**Figure 4:**
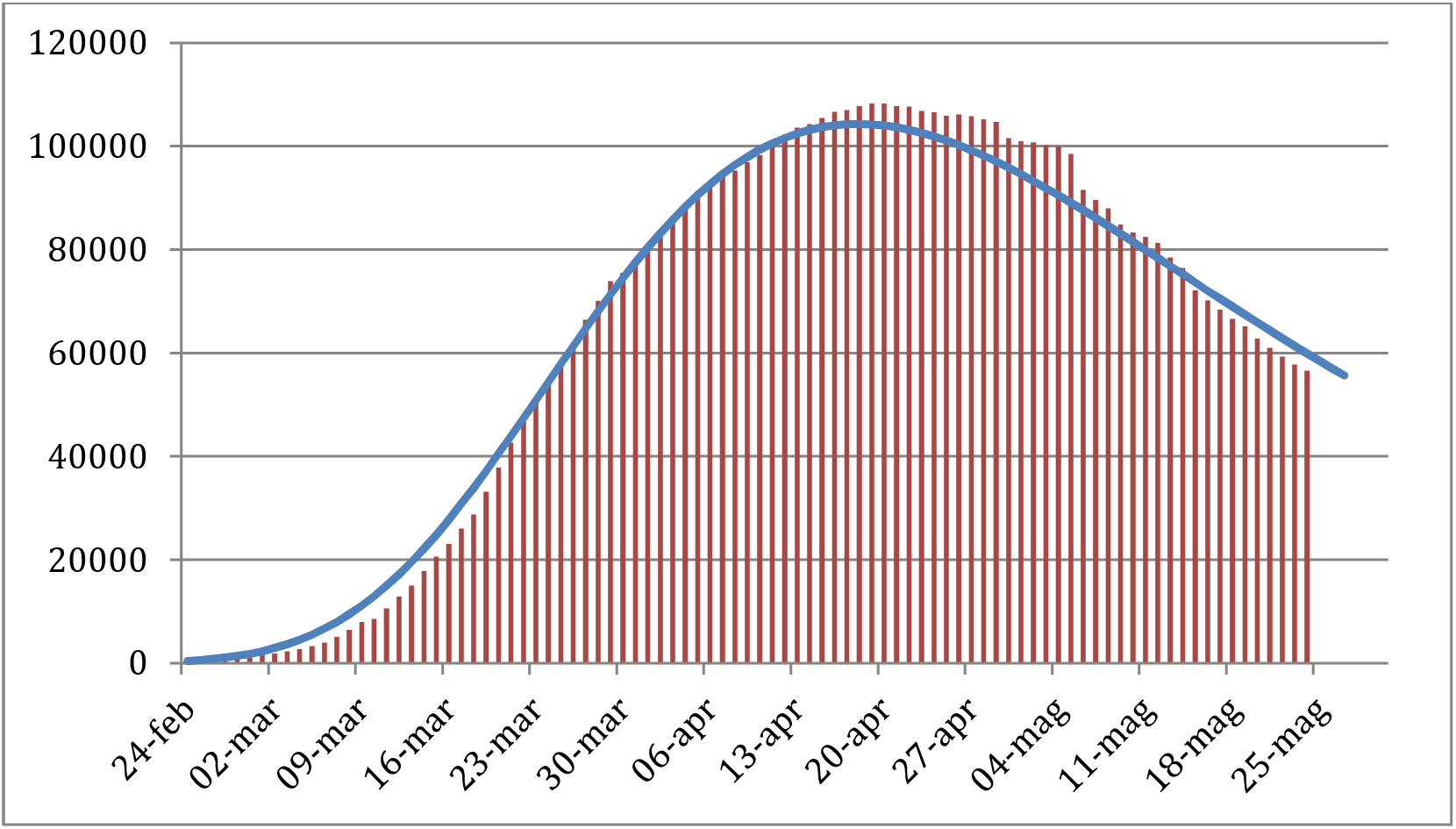
experimental [7] and theoretical curves for *I*(*t*)

By using the same parameters, the number of daily new cases has been calculated from the following equation:

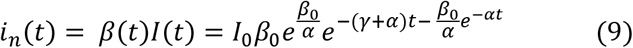

Also *i*_*n*_(*t*) is of the “double exponential” type. Next figure 5 shows an acceptable fit of the experimental data with Equation (9).

**Figure 5:**
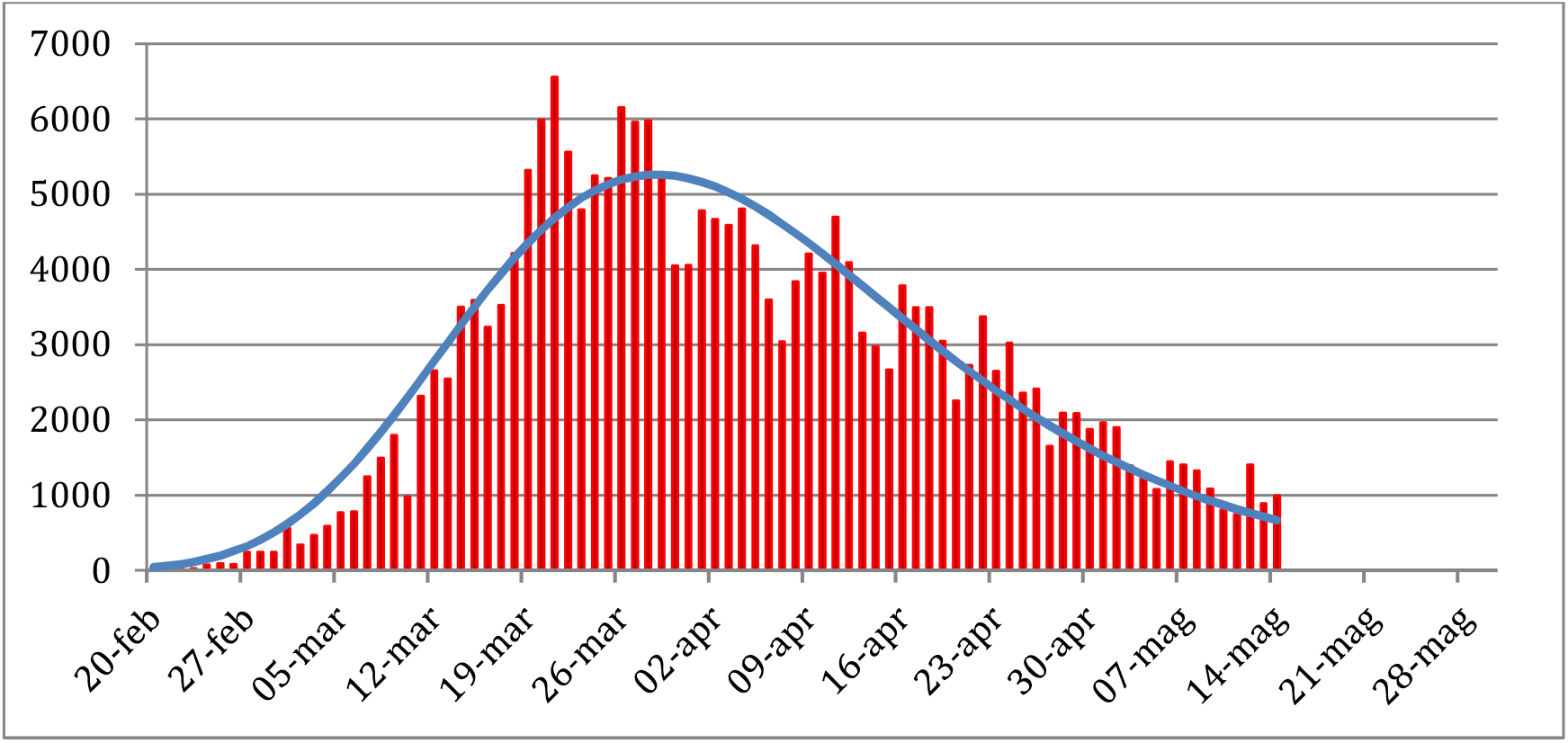
experimental [7] and theoretical curves for *i*_n_(*t*), the daily new infected.

Finally, by numerically integrating Equation (9), we get the total number of infected during the epidemic, that is shown in figure 6:

**Figure 6:**
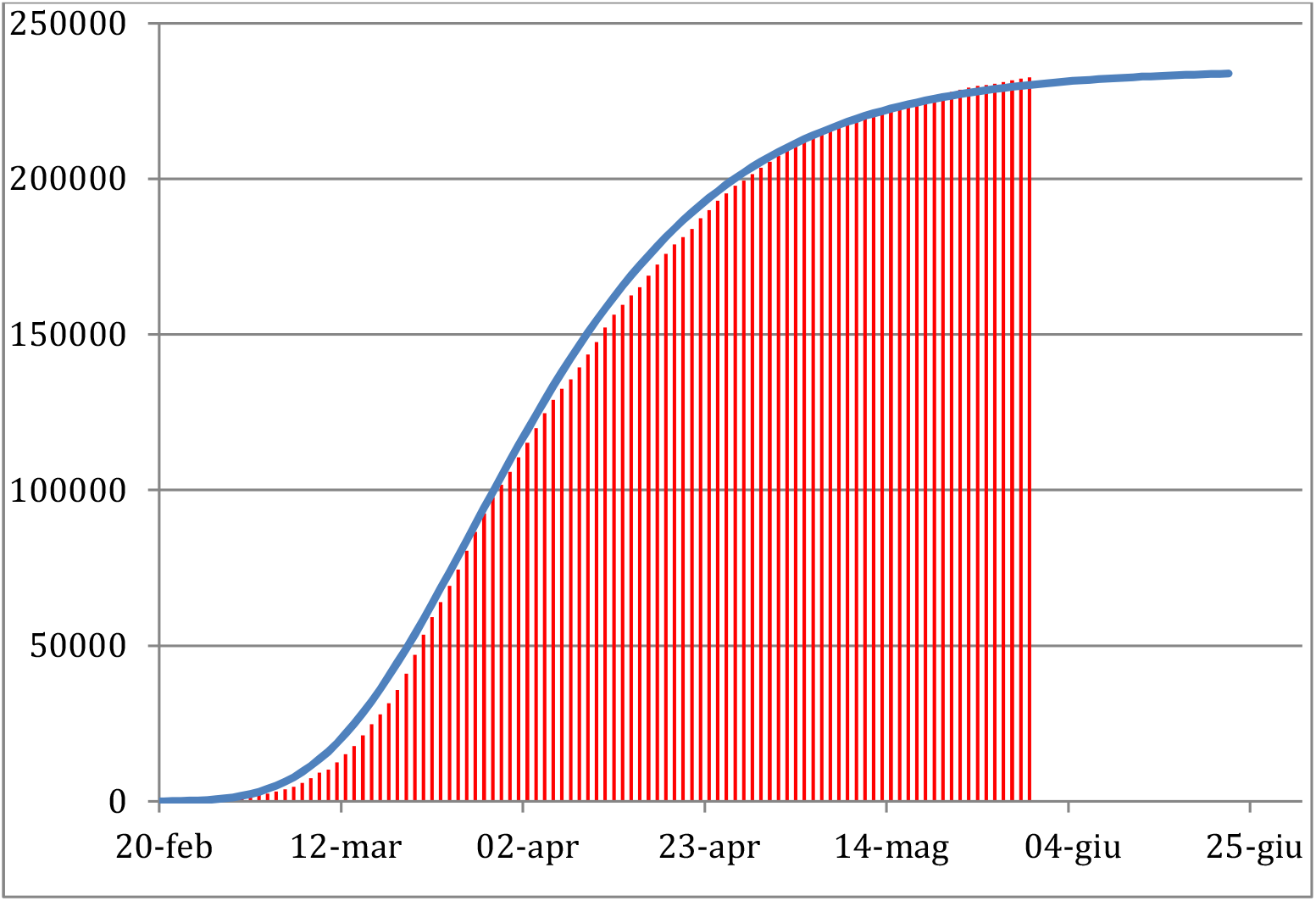
experimental [7] and theoretical curves for *I*_*total*_ (t), the total infected vs. time.

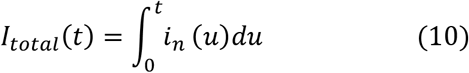

The asymptotic value can be calculated by integrating Equation (10) for *t* → ∞, i.e.:

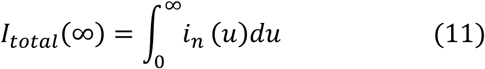

Putting (9) in (11) we get:

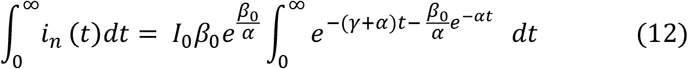

The integral is resolved making reference to the formulas (3.331) in [15] and it can be expressed in terms on the *Incomplete Gamma Function*, defined, according to Nielsen notation [16]:

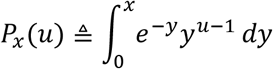

After same calculation we get:

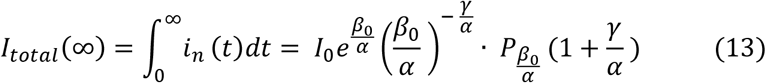

All the parameters are known (*β*_0_ = 0,45, *α* = 0,046, *λ* = 0,03, *I*_0_ = 65) and the asymptotic value given by (13) is around 234.000 total infected. It will be reached in the month of June in Italy.

## Conclusions

On the basis of the classical SIR model, a closed form solution of the differential equations governing the epidemic dynamics has been found. The assumptions made to find the solution are *S/N* ≅ 1, i.e. most of the population still lies on the susceptible compartment at the epidemic end, and *β*(t) has a negative exponential shape. This last conjecture has been verified through the experimental data for Italy.

The solution is of the “double exponential” type and it looks like the Gumbel pdf, i.e. 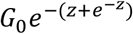. The good fit with experimental data has been finally checked.

## Data Availability

https://datastudio.google.com/reporting/91350339-2c97-49b5-92b8965996530f00/page/RdlHB

## Notes

### Competing Interest Statement

The authors have declared no competing interest.

### Funding Statement

No funding

### Author Declarations

IRB/oversight is not needed for this paper

